# Ursodeoxycholic acid for trans intestinal cholesterol excretion stimulation: a randomized placebo controlled cross-over study

**DOI:** 10.1101/2024.02.22.24303117

**Authors:** R.F. Oostveen, Y. Kaiser, M.L. Hartgers, E.C.E. Meessen, A. Grefhorst, G.K. Hovingh, F. Kuipers, E.S.G. Stroes, A.K. Groen, L.F. Reeskamp

## Abstract

**Objective:** The Trans Intestinal Cholesterol Excretion (TICE) pathway is a potential therapeutic target to reduce plasma low-density lipoprotein (LDL) cholesterol levels. TICE encompasses the direct excretion of cholesterol by enterocytes into feces. In mice TICE has been shown to be stimulated by a hydrophilic bile acid pool, resulting in increased fecal neutral sterols (FNS) loss and reduced plasma cholesterol levels. We investigated whether treatment with a hydrophilic bile acid – ursodeoxycholic acid (UDCA) - would increase FNS in humans as a proxy for TICE.

**Approach and results:** We performed a randomized double-blind placebo controlled cross-over trial in 20 male participants aged >18 years, with plasma LDL cholesterol levels ≥2.6mmol/L. After a run-in period of ezetimibe 20mg once daily for three weeks, patients were randomized to UDCA 600mg or placebo orally once daily for two weeks. After a three week wash-out, patients underwent the alternate treatment.

At baseline, mean (SD) age, BMI, and plasma LDL cholesterol were 59±11.3 years, 26.4±3.1 kg/m2, and 3.9±0.8 mmol/L, respectively. After UDCA treatment, the plasma bile acid hydrophobicity index was reduced compared to placebo (-115.88% versus +1.85%, p<0.001). The FNS did not change (-5.8% versus +18.8%, p=0.51) and treatment with UDCA increased LDL cholesterol with 0.39 mmol/L (+10.95% versus -3.24%, p=0.002) when compared with placebo.

**Conclusion:** UDCA in combination with ezetimibe increased plasma bile acid hydrophilicity in hypercholesterolaemic subjects, but did not result in increased FNS or decreased LDL cholesterol. This suggests that TICE is not stimulated by an increase in the hydrophilicity of the bile acid pool in humans.

## Introduction

Atherosclerotic cardiovascular disease (ASCVD) remains the leading cause of death worldwide.(1) Cholesterol transported in atherogenic lipoproteins, such as low-density-lipoproteins (LDL), is prone to accumulate in the arterial vessel wall, leading to atherosclerotic plaque formation and an increased risk for cardiovascular events.(2) Reducing plasma LDL cholesterol is therefore the cornerstone of cardiovascular risk reduction.(3)

A thus far untapped therapeutic target for LDL cholesterol lowering is the Trans Intestinal Cholesterol Excretion (TICE) pathway. Initially it was believed that cholesterol could only be excreted into feces after being secreted by the liver into bile. However, half a century ago an experiment by Simmonds and colleagues hinted that cholesterol was also actively secreted in the intestinal lumen via a different pathway.(4) Decades later, the importance of this pathway was elucidated in preclinical studies in mice, demonstrating direct cholesterol excretion into feces by enterocytes.(5,6) This process, dubbed TICE, is the net resultant of intestinal cholesterol absorption and excretion and was shown to account for 35% of cholesterol efflux into feces in healthy humans.(7)

The mechanisms controlling the removal of cholesterol through TICE are only partly understood. TICE is mostly mediated by the (chole)sterol exporter ATP-binding cassettes G5 and G8 (ABCG5/G8) but effective therapeutic agents targeting ABCG5/G8 have not yet been developed.(8) However, it has been shown that bile acids, and especially the hydrophilicity of the bile acid pool, are important determinants for cholesterol excretion via ABCG5/G8 in mice.(5) Moreover, the activation of farnesoid X receptor (FXR), which leads to a more hydrophilic bile acid pool in mice, in combination with ezetimibe results in a tremendous increase of TICE by increasing daily sterol excretion up to 60% of their total body cholesterol content.(5) Interestingly, ursodeoxycholic acid (UDCA), a hydrophilic bile acid, strongly decreased biliary cholesterol secretion without influencing fecal neutral sterol secretion in humans.(9,10) Since TICE is the only currently known alternative pathway to remove excess cholesterol from the body, it is plausible that UDCA actually increased TICE in these trials. Studies from our group have shown that ezetimibe treatment can also increase TICE without influencing biliary cholesterol output.(7)

We hypothesized that UDCA in combination with ezetimibe would additively stimulate TICE in humans, and thus result in a more profound increase in fecal neutral sterols and a stronger reduction in LDL cholesterol than ezetimibe alone. To test this hypothesis we performed a randomized double-blind placebo controlled cross-over trial comparing UDCA to placebo on top of ezetimibe treatment in hypercholesterolemic patients.

## Methods

### Study subjects

This study was conducted at the Amsterdam UMC, a tertiary referral hospital in the Netherlands. Subjects were eligible for study participation if they were male, ≥18 years old, and had plasma LDL cholesterol levels ≥2.6 mmol/L. Exclusion criteria were a body mass index (BMI) <19 or >30 kg/m^2^, known history of diabetes mellitus (type 1 or 2), AST or ALT levels ≥2x upper limit of normal (ULN), a recent history of alcohol or drug abuse, contra-indications for the prescription of study drugs (UDCA, ezetimibe), or history of inflammatory bowel disease (e.g. Crohn’s disease or ulcerative colitis). All participants provided written informed consent. The study protocol was approved by the local medical ethics committee and performed in accordance with the Declaration of Helsinki. The study was registered at the Dutch Trial Register (NL6932).

### Study design

This study was a randomized double-blind placebo controlled cross-over trial (**Figure 1**). The study consisted of five study visits with two treatment periods (UDCA or placebo on top of ezetimibe). Participants underwent a screening visit and started ezetimibe 20mg orally once daily after meeting all eligibility criteria. After three weeks of ezetimibe run-in, a baseline visit for the first treatment period was planned. Patients then started with UDCA 600mg orally once daily or matching placebo once daily for two weeks after which an outcome visit for the first treatment period was planned. The study included a three week wash-out period between the first treatment period and the start of the second treatment period, in which the subjects continued using ezetimibe monotherapy. The participants switched to the other treatment arm (from UDCA to placebo or vice versa) in the second treatment period which also consisted of a baseline visit and an outcome visit. Both the investigator and the participant were blinded for treatments sequence allocation and laboratory results obtained after the ezetimibe run-in period. Treatment allocation sequence was determined using a variable randomization block model.

**Figure 1:**
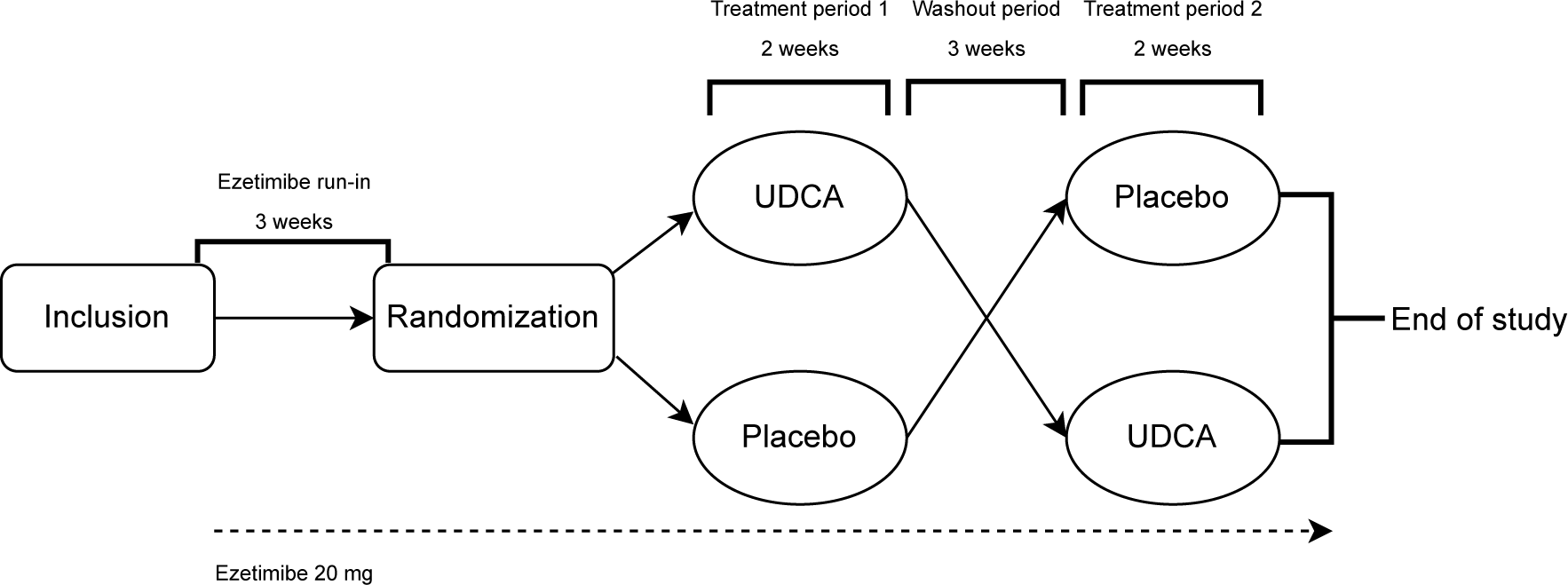
Study-design. UDCA, ursodeoxycholic acid.

### Laboratory analyses

At every visit fasting blood samples were obtained and analyzed for total cholesterol, high-density lipoprotein (HDL) cholesterol, triglycerides, apolipoprotein B100 (apoB), apolipoprotein A1 (apoA1) and lipoprotein (a) (Lp(a)). Lp(a) was measured via an isoform independent, second-generation assay (Roche Diagnostics, Mannheim, Germany). LDL cholesterol was calculated with the Friedewald formula.(11) Fibroblast growth factor 19 (12) (FGF19) was measured in plasma using a Human FGF19 Quantikine ELISA kit (DF 1900, R&D Systems, Minneapolis, MN, USA) according to the manufacturer’s instructions. FGF19 was undetectable below 15.6 pg/mL. In order to avoid under- or overestimation of the effect size missing values were replaced by7.8 pg/mL. Feces samples, were weighed, diluted and homogenized prior to quantification of bile acids and neutral sterols. Plasma bile acid species were quantified by liquid chromatography–tandem mass spectrometry (LC/MS/MS) procedures using a Nexera X2 Ultra High Performance Liquid Chromatography system (SHIMADZU, Kyoto, Japan), connected to a SCIEX QTRAP 4500 MD triple quadrupole mass spectrometer (SCIEX, Framingham, MA, USA) (UHPLC-MS/MS) as previously described.(13) In weighted stool samples, concentrations of bile acids and the neutral sterols cholesterol, dihydrocholesterol and coprostanol were measured.(7)

### Hydrophobicity index

To calculate the total plasma bile acid hydrophobicity index (HI) we used the previously published HI for each individual bile acid(14), multiplied by its relative concentration (formula reported in Supplementary Methods). The sum of the resulting HI’s represents the total plasma bile acid HI.

### Primary and secondary outcomes

The primary outcome was percentage change in FNS concentrations after UDCA treatment compared to placebo treatment. Secondary outcomes were the effects of UDCA treatments on the plasma bile acid hydrophilicity index, as well as plasma LDL cholesterol, total cholesterol, HDL cholesterol, triglycerides, Lp(a), apoB, apoA1 and FGF19 levels.

### Statistical analyses

This was a proof-of-concept study and the effect of combination therapy of UDCA and ezetimibe on FNS excretion has not been studied before. Based on a previous study, which showed that total cholesterol excretion after administration of ezetimibe was 1939 mg/day with an SD of 658 mg/day (7), a sample size of 20 subjects has more than 80% power to detect a difference in means of 20% after treatment with UDCA using a crossover study design. All data are expressed as mean ± standard deviation, median [interquartile range] or number (percentage) when appropriate. Considering the half-life of UDCA, we did not expect carry-over nor period effects to be a factor, thus changes in normally distributed data were assessed using a paired t-test, whereas those between non-normally distributed data were assessed using the paired samples Wilcoxon Signed-Rank test. Finally, we tested correlations between LDL cholesterol, FNS, and plasma bile acid hydrophilicity using the Pearson correlation coefficient or Spearman’s rank correlation coefficient for normally and non-normally distributed data respectively. Statistical significance was defined as p<0.05. All statistical analyses were performed in R version 4.0.2 (R Foundation, Vienna, Austria).

## Results

A total of 20 male subjects participated in this study between October 2018 and October 2019 at the Amsterdam UMC. One participant was excluded from all analysis regarding fecal outcomes, because of missing data at the final study visit. No patients were on statin therapy at the start of the study, and no patient had a history of ASCVD. Mean age, BMI, and LDL cholesterol were 59 ± 11.3 years, 26.4 ± 3.1 kg/m^2^, and 3.9 ± 0.8 mmol/L, respectively (**Table 1**). Median Lp(a) and mean HbA1c were 183 [119.75, 404.75] mg/L and 31.7 ± 31.04 mmol/mol, respectively.

**Table 1:** Characteristics of participants at baseline.

|  | **Study participants** |
| --- | --- |
| **n** | 20 |
| **Age, years** | 59.4 ± 11.3 |
| **Sex, male** | 20 (100%) |
| **BMI, kg/m<sup>2</sup>** | 26 ± 3 |
| **Systolic blood pressure, mmHg** | 137 ± 18 |
| **Diastolic blood pressure, mmHg** | 80 ± 11 |
| **Total cholesterol, mmol/L** | 6.0 ± 1.0 |
| **LDL cholesterol, mmol/L** | 3.9 ± 0.8 |
| **HDL cholesterol, mmol/L** | 1.3 ± 0.3 |
| **Triglycerides, mmol/L** | 1.5 [1.1, 2.0] |
| **Lipoprotein (a), mg/L** | 183. [120, 405] |
| **Billirubine, U/L** | 9 ± 4 |
| **Alkaline phosphatase, U/L** | 72 ± 18 |
| **gGT, U/L** | 33 ± 16 |
| **ALT, U/L** | 27 ± 12 |
| **AST, U/L** | 15 ± 38 |
| **HbA1c, mmol/mol** | 32 ± 31 |
| **Lipid lowering therapy** | 0 (0%) |

Numbers present n (%), mean +/- SD, or median [interquartile range]. BMI, body mass index; LDL, low-density lipoprotein; HDL, high-density lipoprotein; gGT, gamma-glutamyl transferase; ALT, alanine aminotransferase; AST, aspartate aminotransferase.

The FNS levels were not affected by UDCA nor placebo treatment and showed no statistical difference between the two treatments (-1.41 nmol/mg versus 4.42 nmol/mg, p=0.54) (**Figure 2**). The mean increase of fecal coprostanol and cholestanol content after UDCA treatment was not different compared to placebo (2.77 nmol/mg versus 2.37 nmol/mg, p=0.95, and -4.18 nmol/mg versus 2.05 nmol/mg, p=0.19 respectively) (**Table 2**).

**Figure 2.**
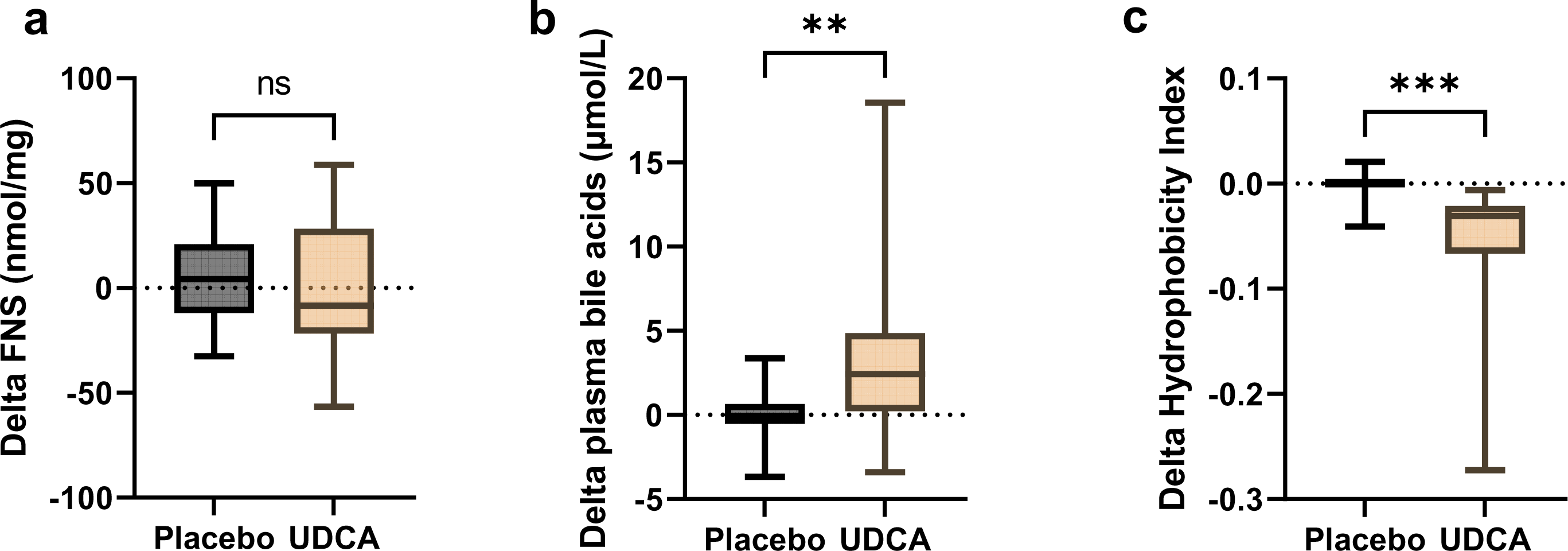
Effect of UDCA on FNS, plasma bile acids and hydrophobicity index. FNS, fecal neutral sterols; UDCA, ursodeoxycholic acid.

**Table 2:**
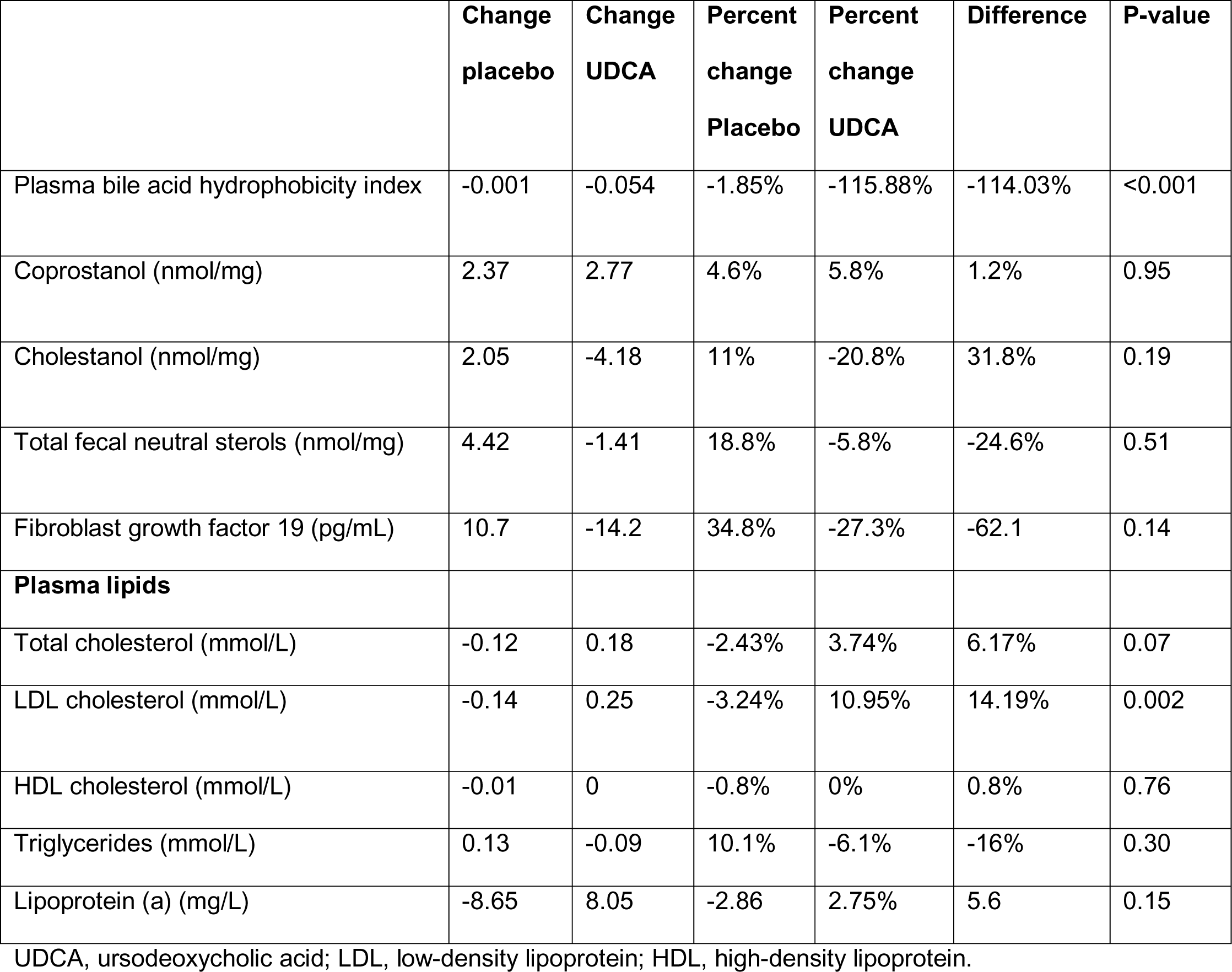
Efficacy Results.

UDCA treatment significantly increased total plasma bile acid concentration from 2.9 μmol/L to 6.7 μmol/L compared to a 2.0 μmol/L to 1.9 μmol/L change in placebo (p<0.01) (**Figure 2**). This change coincided with a decrease in mean plasma bile acid hydrophobicity index compared to placebo (-0.053 versus - 0.001, p<0.001) (**Figure 2**), -115.88% and +1.85% respectively. The plasma bile acid hydrophobicity index did not correlate with FNS (ρ=0.1, p=0.41, **Figure 3**).

**Figure 3.**
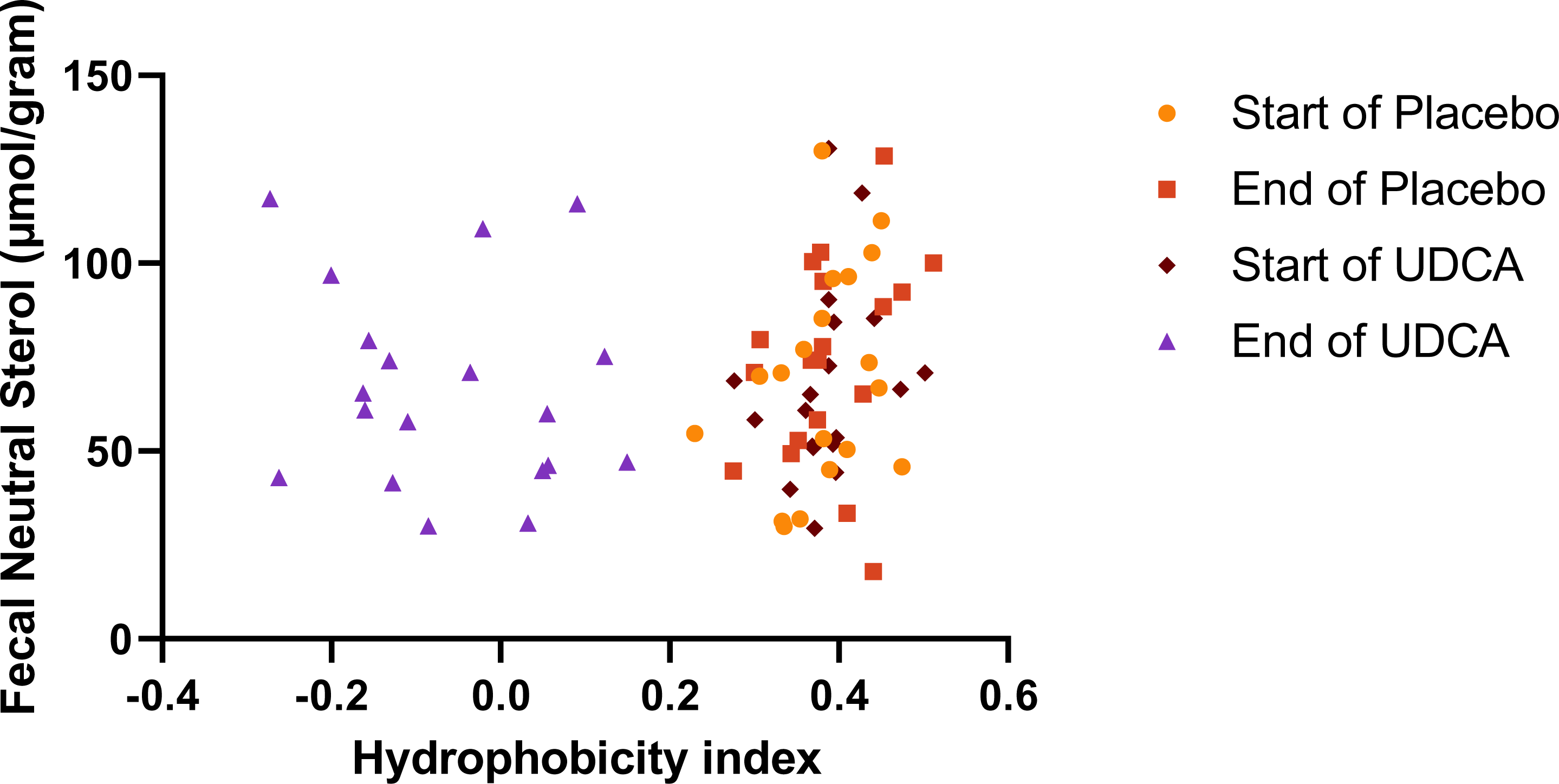
Correlation FNS – Hydrophobicity. UDCA, ursodeoxycholic acid

The mean changes in total plasma cholesterol and LDL cholesterol levels on UDCA treatment were +0.18 mmol/L and +0.25 mmol/L, respectively, compared to -0.12 mmol/L and -0.14 mmol/L, respectively, in the placebo group. Compared to placebo, the increase in plasma LDL cholesterol levels upon UDCA was significantly higher than in the placebo subjects (p=0.002) (**Figure 4**). No significant changes upon UDCA treatment were observed in plasma HDL cholesterol, triglycerides and Lp(a) levels (**Table 2**, **Figure 4**).

**Figure 4.**
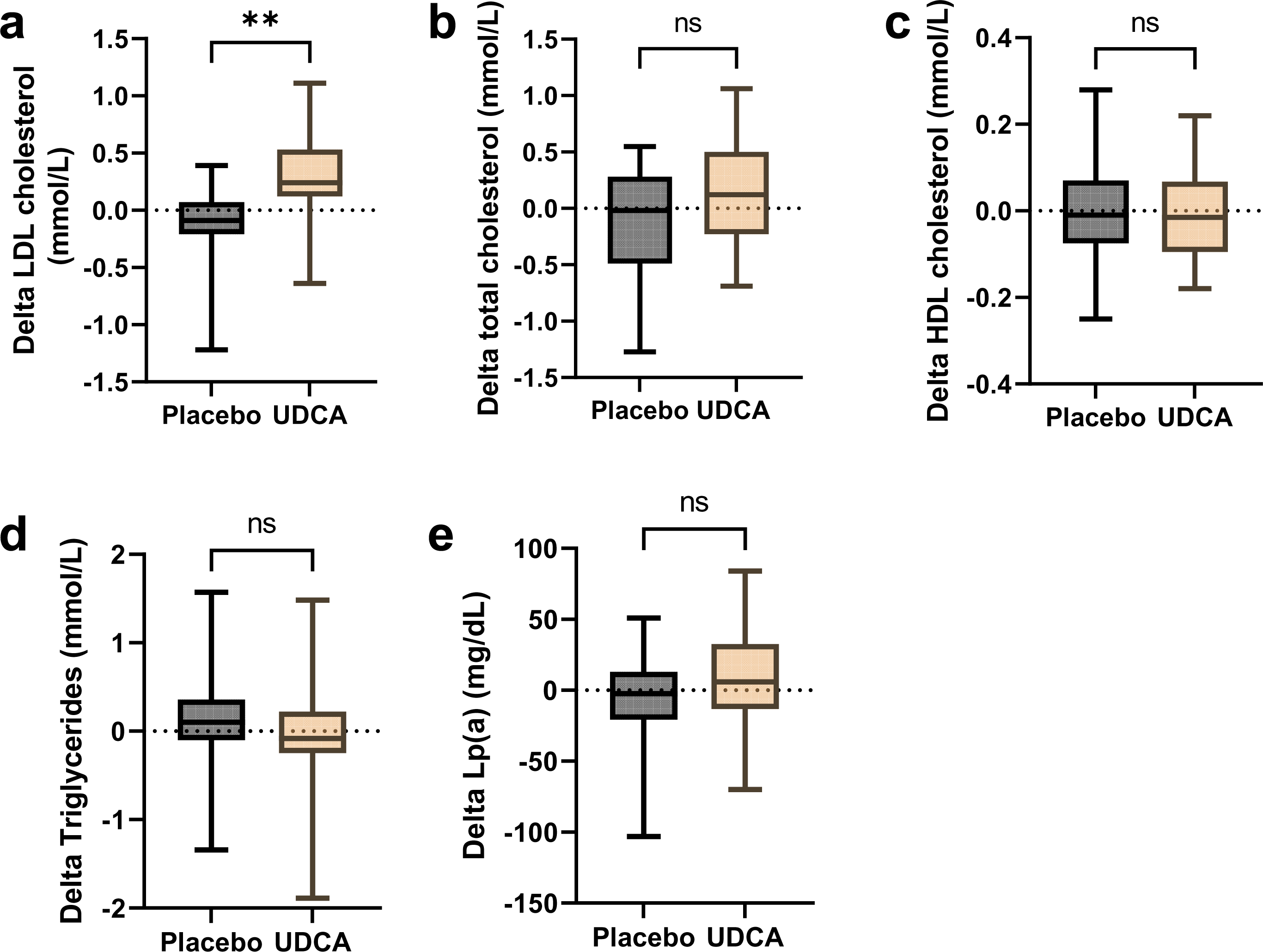
Lipid changes. UDCA, ursodeoxycholic acid; LDL, low-density lipoprotein; HDL, high-density lipoprotein; Lp(a), lipoprotein (a).

Since TICE is in part regulated by FXR through FGF19 in mice, we examined the effect of UDCA treatment on plasma FGF19 levels. The plasma FGF19 level was below the detectable limit of 15.6 pg/mL in 15 out of the total 76 samples measured. FGF19 measurements revealed an outlier with consistently undetectable levels at all timepoints, prompting its exclusion from subsequent analyses. There was a non-significant decrease in FGF19 after treatment with UDCA compared to placebo (-14.2 pg/mL versus +10.7 pg/mL, respectively). These values correspond to percentual changes of -27.3% and +34.8% (p=0.14), respectively There was no correlation between a change in FGF19 and a change in LDL cholesterol (r=0.1, p=0.70), nor was there a correlation between FGF19 and HI after two weeks of UDCA treatment (r=0.26, p=0.28) (**Figure 5**).

**Figure 5.**
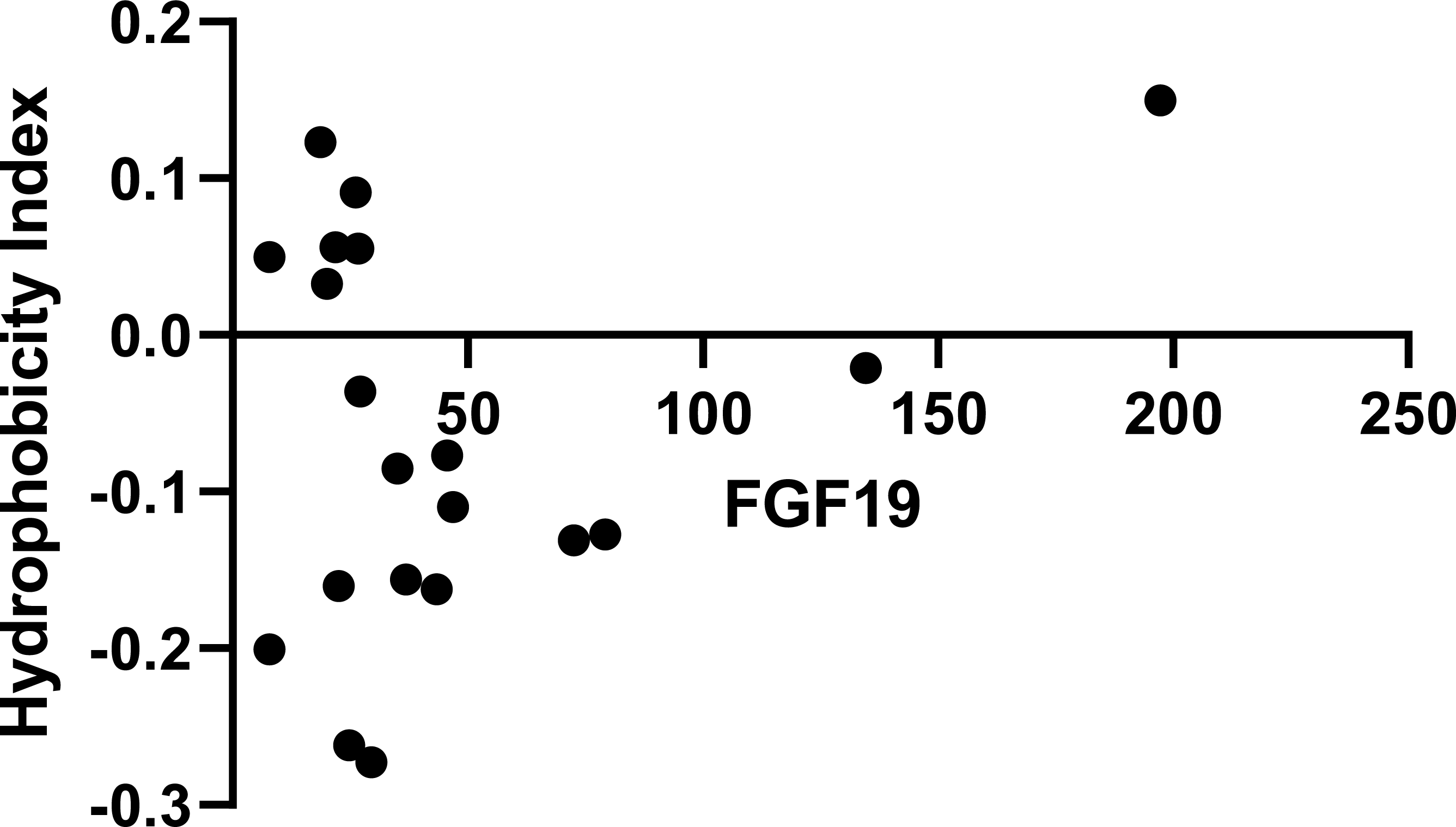
Correlation FGF19 – Hydrophobicity index. FGF19, fibroblast growth factor 19.

## Discussion

In this randomized placebo-controlled cross-over trial we tested the hypothesis that treatment with UDCA results in a more hydrophilic bile acid pool in hypercholesterolemic subjects that would thereby increase FNS as a proxy for TICE. Despite an increase in hydrophilicity of circulating bile acids, FNS was not increased compared to placebo after 3 weeks of UDCA treatment. Surprisingly, plasma LDL cholesterol levels were significantly increased after treatment with UDCA.

As shown in previous studies, treatment with UDCA resulted in a more hydrophilic bile acid pool.(15) However, in contrast to studies in mice, in which an increase in bile acid hydrophilicity resulted in increased FNS excretion (5), we were unable to confirm this in hypercholesterolemic humans. Moreover, we did not observe any correlation between the plasma bile acid hydrophobicity index and FNS. Several explanations for this discrepancy can be considered. First, there may be fundamental differences between the human and murine ABCG5/G8, the responsible transmembrane transporter of cholesterol in enterocytes, for example in their affinity towards hydrophilic bile acids. Second, it is conceivable that the correlation between HI and FNS may only become apparent in humans when a certain HI threshold is met. The lowest HI that was reached in this study was -0.27, whereas in mice the correlation between HI and FNS did not become apparent until a HI below -0.4.(5). It is important to note that the composition of murine bile acid pool fundamentally differs from that of humans due to the presence of mouse/rat specific bile acid species, i.e., the α-, β-, and ω-muricholic acids. These very hydrophilic bile acids give rise to a very low HI of the murine bile acid pool that potentially has a stronger stimulatory effect on TICE compared to UDCA in humans.(14) Lastly, UDCA has been shown to have antagonistic effects on FXR-controlled pathways in patients with morbid obesity, resulting in a reduction of FGF19 levels and thus less FNS excretion.(16) It is therefore possible that the negative results of our study are the result of the antagonistic effects of UDCA on FXR that dampens its potential beneficial effect on FNS.

Surprisingly, UDCA treatment in combination with ezetimibe resulted in a 14.19% increase in plasma LDL cholesterol levels (p=0.005) and a 6.17% non-significant increase in plasma total cholesterol levels (p=0.07) when compared to placebo. This effect was observed on top of the expected ezetimibe-induced reduction of LDL cholesterol (21%) during the run-in period. This UDCA-related increase in plasma cholesterol has not been reported in previous trials with UDCA. For example, a trial on healthy subjects showed that plasma LDL cholesterol levels remained unchanged on UDCA treatment.(17) Additionally, a recent meta-analysis concluded that UDCA treatment may significantly reduce plasma LDL cholesterol levels in patients with primary biliary cholangitis.(18) It has been suggested that this may be due to inhibition of hydroxymethylglutaryl-co-enzyme A reductase (19) or a decrease in dietary cholesterol absorption.(20) However, both hypotheses remain to be evaluated. Since FNS remained unchanged in our participants, it is unlikely that the observed increase in LDL cholesterol in our study was due to altered intestinal sterol handling. Moreover, plasma FGF19 was unchanged, indicating that activation of intestinal FXR was not impaired.

## Conclusion

This double-blinded randomized placebo-controlled cross-over trial demonstrated that an increased plasma bile acid hydrophilicity achieved with UDCA therapy in humans does not result in an increase in FNS excretion nor in reduced plasma LDL cholesterol levels. Thus, UDCA is unlikely to be a potential modulator of TICE in humans. Nonetheless, TICE still holds potential as a therapeutic target in reducing plasma LDL cholesterol. More insight in the mechanisms involved in TICE is needed before therapeutic methods to control TICE can be successfully be explored.

## Data Availability

All data produced in the present study are available upon reasonable request to the authors

