## Supplemental Methods for "Ursodeoxycholic acid for trans intestinal cholesterol excretion stimulation: a randomized placebo controlled cross-over study"

*Hydrophobicity index*

To calculate the total plasma bile acid hydrophobicity index (HI) we used the previously published HI for each individual bile acid[1] and multiplied them by the relative concentration of each bile acid.

*Formula:*

Total plasma bile acid HI=

((bile acid 1 concentration * bile acid 1 HI)/ total bile acid amount)

+ (bile acid 2 concentration * bile acid 2 HI)/ total bile acid amount)

+ …

+ (bile acid 14 concentration * bile acid 14 HI)/ total bile acid amount))
